# Test Development with Content Validity in Pediatric Hematology

**DOI:** 10.1101/2023.01.21.23284870

**Authors:** Beeling M. Armijo, Ghada A. Abusin, Clarence D. Kreiter, Patrick B. Barlow, Steven R. Lentz

**Author notes:** Corresponding Author: Steven R. Lentz, MD, PhD, Department of Internal Medicine, The University of Iowa Carver College of Medicine, 200 Hawkins Drive C21 GH, Iowa City, IA 52242.

## Abstract

Content validity is often lacking in tools for the assessment of competency in medical specialty and subspecialty trainees. The primary aim of this study was to develop a protocol for early test assessment in pediatric hematology using iterative rounds of expert evaluation in the context of explicit knowledge goals. We used the pediatric hematology content of the American Board of Pediatrics as a test case for assessment protocol development. We found that the use of expert input and three rounds of test development increased test content validity. The instrument developed here has the potential to improve assessment of competency in pediatric hematology. This approach also may be useful in the development of competency assessment tools for other medical subspecialties.

## Introduction

Pediatric residency programs have undergone major paradigm shifts over the last decade largely due to advancements in medicine and duty hour restrictions. As the delivery of health care and medicine evolves, subspecialty clinician educators are faced with the challenge of maximizing teaching opportunities in the face of greater time constraints. The complexity of this challenge is increased by the added necessity of adapting teaching methods to appropriately address the learning styles of each unique generation of trainees. The United States Accreditation Council on Graduate Medical Education’s (ACGME) stringent educational requirements for pediatric training programs are poured on top of an already steep embankment of clinical and/or research responsibilities of an academic subspecialty educator.^1^ To help guide teaching efforts the American Board of Pediatrics (ABP) publishes regularly updated subspecialty content specifications, which outline the information residents are expected to master in each respective field.^2-4^ The current literature that evaluates testing in subspecialty education is very limited.^1^

Current literature describing subspecialty educational tools and interventions lacks in-depth review and analysis of the initial steps in planning and development of such tools. These steps are critical because they ensure the reliability and validity of assessment tools.^5-9^ The primary aim of this study is to develop a protocol for early test assessment in pediatric hematology using iterative rounds of expert evaluation of assessment tools in the context of explicit knowledge goals. This approach allows an explicit evaluation of content validity, a crucial step in assessment development that is often lacking in subspecialty assessment evaluation. Content validity refers to the independent assessment of test material to determine whether the content sampled matches the learning domain of interest.^10^ We used the pediatric hematology content of the ABP as a test case for our assessment protocol.

## Methods

We first conducted a needs assessment to determine if a current resident educational evaluation tool in pediatric hematology existed and the extent of its utility and application. The needs assessment included interviews with our institution’s residency program educational director, pediatric hematology faculty, and trainees. We found that there was no current pediatric hematology subspecialty educational evaluation tool for trainees. Yearly in-training examinations (ITE) are used as a self-assessment instrument for residents. Residency programs commonly use ITE scores to gauge a resident’s likelihood of passing the ABP General Pediatric Certifying Exam.^11^

Given the results of our needs assessment, we endeavored to develop an assessment instrument. We started by reviewing the content specifications published in January 2016 by the ABP for the General Pediatric Certifying Exam.^2^ The content specifications comprise a list of topics in all areas of pediatrics that residents are expected to master as evidence of their competency as they go on to practice medicine. A total of 113 content specifications were listed for blood and neoplastic disorders; 86 (76%) of these items pertain to topics in non-malignant hematology. These 86 items were sub-divided into six sub-categories: general aspects, erythrocyte disorders, leukocyte disorders, platelet disorders, pancytopenia and coagulation disorders. The six sub-categories may be regarded as a fixed facet within the larger domain, with items primarily nested within sub-categories. A facet represents a set of measurement conditions that are a possible source of variation in our study. The six sub-categories are fixed facets because they contain only the conditions of interest and are not a sample from a larger population of categories. Each item nested within sub-categories serves to clarify what information residents are expected to master.

We drafted 40 multiple-choice questions to test trainees’ knowledge of hematology. These questions were modeled after the ABP’s list of specifications for non-malignant hematology (Table 1). Assessment questions were crafted with the help of experienced faculty in pediatric hematology-oncology with a background of formal training in medical education, as well as expert faculty in medical education and psychometrics. The resulting hematology test was first administered to two-experienced faculty in pediatric hematology oncology at separate academic institutions. Revisions to the test questions and answer choices were made based on feedback received from these faculty.

**Table 1.** Frequency and characteristics of changes in test question and answer choice sets

| Number of revisions<sup>1</sup>, (N) | Item<sup>2</sup> | ABP Content specification<sup>3</sup> | Summary of comments/recommendations<sup>4</sup> |
| --- | --- | --- | --- |
| 3 | Disorder of erythrocyte production | Erythrocyte disorders | More background and description of anemia; question does not clearly address content specification |
| 3 | Iron deficiency anemia | Erythrocyte disorders | More background and description of anemia; more than one correct answer choice |
| 3 | Absorption of iron | Erythrocyte disorders | Exceeded goal level of desired knowledge for learner; more recent literature conflicts with answer choices; clarify answer choices |
| 2 | Fanconi anemia | Platelet disorder | Exceeded goal level of desired knowledge for learner; more than one correct answer choice |
| 2 | Thrombocytopenia and bruising | General aspects of blood disorders | More background information; clarify differentiation from injuries related to falls versus other causes of thrombocytopenia |
| 2 | B-12 deficiency | Erythrocyte disorders | More background and description of anemia |
| 2 | Correction of iron deficiency anemia | Erythrocyte disorders | Answer choices not appropriate for level of the learner; edit answer choices to test broader scope of knowledge |

<sup>1</sup>All reviewers agreed that item was high quality = 0, no revisions were made.
<sup>2</sup> Topics covered in test questions and answer choice sets.
<sup>3</sup> ABP content specification published in January 2016.<sup>2</sup>
<sup>4</sup> Expert reviewer comments and recommendations following review of test questions/answer choice sets.

| Number of revisions<sup>1</sup>, (N) | Item<sup>2</sup> | ABP Content specification<sup>3</sup> | Summary of comments/recommendations<sup>4</sup> |
| --- | --- | --- | --- |
| 2 | Vaso-occlusive crisis in sickle cell disease | Erythrocyte disorders | Clinical vignette did not adequately describe disease phenotype; more than one correct answer choice available |
| 2 | ABO incompatibility | Erythrocyte disorders | Exceeded goal level of desired knowledge for learner; more background information necessary; background information confusing |
| 2 | Diamond-Blackfan syndrome | Erythrocyte disorders | More background information necessary; more than one correct answer choice |
| 2 | ITP | Platelet disorder | Controversial choice of management; edit wording used in answer choice |
| 2 | Neonatal thrombocytopenia | Platelet disorder | More than one right answer to choose from; answer choice confusing |
| 2 | Catheter related thrombosis | Coagulation disorders | Vignette and corresponding question are not cohesive; answer choices are too similar |
| 1 | Diagnostic evaluation of multiple pancytopenias | Platelet disorder | Clarify question stem |
| 1 | Folate deficiency | Erythrocyte disorders | Exceeded goal level of desired knowledge for learner |
| 1 | Lead poisoning | Erythrocyte disorders | Did not address content specification |

| Number of revisions<sup>1</sup>, (N) | Item<sup>2</sup> | ABP Content specification<sup>3</sup> | Summary of comments/recommendations<sup>4</sup> |
| --- | --- | --- | --- |
| 1 | Hereditary spherocytosis | Erythrocyte disorders | Background information confusing |
| 1 | Management of thalassemia major | Erythrocyte disorders | More than one right answer to choose from; answer choice confusing |
| 1 | G6PD deficiency | Erythrocyte disorders | Clinical vignette did not adequately describe disease phenotype |
| 1 | Blood product transfusion | General aspects of blood disorders | Identify as historical question to understand current standard of care |
| 1 | ITP | Platelet disorder | Clarify answer choices |
| 1 | Wiskott-Aldrich syndrome | Platelet disorder | Did not address content specification |
| 1 | Glanzmann syndrome | Platelet disorder | More background information necessary |
| 1 | Infection risks associated with neutropenia | Leukocyte disorders | Background information confusing |
| 1 | Chronic granulomatous disease | Leukocyte disorders | More background information necessary |
| 1 | Severe congenital neutropenia | Coagulation disorders | Confusing answer choice |
| 1 | Diagnostic evaluation of bleeding diatheses | Coagulation disorders | Controversial choice for work-up |

| Number of revisions<sup>1</sup>, (N) | Item<sup>2</sup> | ABP Content specification<sup>3</sup> | Summary of comments/recommendations<sup>4</sup> |
| --- | --- | --- | --- |
| 1 | Disseminated intravascular coagulopathy | Coagulation disorders | Background information confusing |
| 1 | FVIII deficiency | Coagulation disorders | Background information confusing |
| 1 | von Willebrand disease | Coagulation disorders | Clinical vignette is confusing |
| 0 | Physiologic anemia of infancy | General aspects of blood disorders |  |
| 0 | Diagnosis of sickle cell disease at birth | Erythrocyte disorders |  |
| 0 | Prophylactic measures for sickle cell disease | Erythrocyte disorders |  |
| 0 | Prophylactic measures for hereditary spherocytosis |  |  |
| 0 | Transient erythroblastopenia of childhood (TEC) |  |  |
| 0 | Causes of pancytopenia | Platelet disorder |  |
| 0 | Causes of thrombocytopenia | Platelet disorder |  |
| 0 | TAR syndrome | Platelet disorder |  |

| Number of revisions<sup>1</sup>, (N) | Item<sup>2</sup> | ABP Content specification<sup>3</sup> | Summary of comments/ recommendations<sup>4</sup> |
| --- | --- | --- | --- |
| 0 | Management of leukocyte disorder | Leukocyte disorders |  |
| 0 | Vitamin k deficiency | Coagulation disorders |  |

Next, four experts in pediatric hematology from four separate academic institutions volunteered to participate as test reviewers in our study. Each expert included in our study is a recognized expert in pediatric hematology. Expert reviewers comprised greater than 30 years of combined knowledge and experience in pediatric hematology.

An explicit set of scoring guidelines, referred to as a scoring rubric, was given to each expert reviewer. The scoring rubric was used to ensure that reviewers scored questions and answer choices consistently and in a manner comparable to other reviewers to enhance the reliability of the rating data. Reviewers were given the choice to score each test question or set of answer choices as a “high quality item = 3” no changes recommended; “adequate item = 2” but needs improvement/editing; or “inadequate item =1” discard question. Reviewers were asked to provide comments explaining their decision if a test question or set of answer choices was scored a two (i.e. adequate item) or a one (i.e. inadequate item). After each separate review, revisions to test questions were made in response to reviewer comments.

Descriptive statistics were used to summarize the results of expert evaluations. Absolute agreement in the form of percent agreement was used to report the results of expert scoring. An adjustment for chance agreement among judges was not informative for our study due to the expertise among judges (e.g., when four experts agree, there is a very little possibility that their agreement is due to chance).

## Results

Expert judges reviewed all items of our hematology test three separate times until 100% agreement was reached. After the first evaluation, we found 30% of the test questions/answer choice sets were rated as high quality items (Table 1). Seventy percent of the questions/answer choice sets were revised and/or changed based on expert reviewer comments and recommendations from the first evaluation. After reviewers evaluated all 40 questions/answer choice sets (included unchanged and revised material) a second time, 78% of the test items were found to be high quality amongst all judges. The remaining 22% of test questions/answer choice sets that were not found to be high quality were revised based on expert judge comments and recommendations. The third and final review of test questions/answer choice sets by expert judges only included items that were revised after the second evaluation. Expert judges were not asked to review all 40-test questions/answer choice sets after the second evaluation due to the high time burden this would place on our reviewers. Expert judges reported 100% of the test questions/answer choice sets that were revised following the second evaluation to be high quality items on the third evaluation.

## Discussion

The goal of our study was to assess the efficacy of applying a rigorous iterative approach to test development and learning from medical education theory to the issue of resident ABP competence in pediatric hematology. We found that the use of expert input and three rounds of test development increased test content validity. This approach also may be useful in assessment development for other subspecialties.^12^

An important outcome of our study is that the rated item quality increased with each round of review, with eventual rater consensus regarding item quality. Taken together, these results provide evidence for content validity. Failure to provide adequate evidence for content validity undermines the entire purpose of educational assessment. A common practice among academic programs is to use residents’ yearly ITE scores as a surrogate marker to assess their knowledge of hematology.^11^ The ITE was developed by the ABP to assess residents’ overall yearly mastery of a broad spectrum of topics in pediatrics. However, a major problem with using ITE scores to address ABP-level expertise in pediatric subspecialties is the limited ability of the ITE to provide adequate content validity to address the learning domains outlined by the ABP. For example, the number of hematology questions included on the ITE exams from 2014 to 2017 ranged from 4-7 questions. At best seven questions were asked from the 86 possible topics listed by the ABP as essential for residents to master in hematology. The vast majority of topics missing from ABP section on hematology on the ITE render the scores in this particular area not a useful tool to gauge the effect of our teaching as subspecialists. Any inferences or correlations regarding a resident’s knowledge of hematology based on ITE scores are difficult to postulate for this reason.

The instrument developed here has the potential to improve assessment of competency in pediatric hematology but future work to further develop the tool is needed. In particular, research to evaluate what topics expert reviewers found missing or not adequately represented within the list of content specifications published by the ABP is still needed. Our study utilized an established procedure for test development in medical education and applied it to an environment that is largely unfamiliar with its use in resident education.^12^ It is not difficult to understand how the vast amount of work and collaboration required to construct and administer valid assessment instruments can be daunting amidst a physician’s busy clinical and/or research schedule. However, pediatric hematology and many other subspecialities are based primarily in academic medicine. As such, these fields of medicine will always be dependent on the participation of learners. Developing educational assessment instruments that demonstrate rigorous evidence for content validity is an essential responsibility of every clinician educator.

## Data Availability

All data produced in the present study are available upon reasonable request to the authors

